# Evaluation of Effects on Skin Quality of a *Centella asiatica* Extracellular Vesicle-based Skin Care Formulation: A 28-Day Facial Skin Quality Study

**DOI:** 10.1101/2025.03.04.25323195

**Authors:** Tsong-Min Chang, Chung-Chin Wu, Huey-Chun Huang, Shr-Shiuan Wang, Ching-Hua Chuang, Pei-Lun Kao, Wei-Hsuan Tang, Luke Tzu-Chi Liu, Wei-Yin Qiu, Ivona Percec, Charles Chen, Tsun-Yung Kuo

**Affiliations:** Department of Applied Cosmetology, HungKuang University, Taichung City, Taiwan; Schweitzer Biotech Company, Taipei City, Taiwan; Department of Medical Laboratory Science and Biotechnology, China Medical University, Taichung City, Taiwan; Department of Clinical Application, Center for iPS Cell Research and Application (CiRA), Kyoto University, Kyoto, Japan; Division of Plastic Surgery, Department of Surgery, University of Pennsylvania, Philadelphia, PA, USA; College of Science and Technology, Temple University, Philadelphia, PA, USA

**Keywords:** *Centella asiatica*, Cica, skincare, cosmetics, longevity, extracellular vesicles, skin quality, skin hydration, skin whitening, skin elasticity, wrinkle reduction

## Abstract

Skin damage results from the natural aging process, physical damage, UV light, and environmental pollutants. *Centella asiatica* (Cica) is a traditional medicinal herb known to have therapeutic effects on skin and wound healing. This study aims to evaluate the effects of skincare formulation Kristen Claire Supreme Rejuvenation Essence with Cica EV as the main active ingredient, on facial appearance and skin quality in healthy participants. Twenty healthy participants (4 males and 16 females; average age 36.5) were enrolled and underwent a 24-hour skin patch test on the forearm to assess potential irritation or allergic reactions. The test product was applied twice daily for 28 days. Facial skin quality assessments were conducted before use and at 7, 14, 21, and 28 days of product application. Measured parameters included skin hydration, melanin content, skin elasticity, wrinkle percentage, redness area percentage, and pore percentage. After 28 days of test product treatment, significant improvements were observed in skin hydration and elasticity, wrinkle, redness, and pore distribution, while skin melanin content was reduced significantly. Kristen Claire Supreme Rejuvenation Essence effectively improved skin hydration, elasticity, and texture, and brightened the complexion, making it a promising candidate for skincare applications.

## Introduction

As the largest organ of the human body, skin is the main barrier of entry which protects the internal body from environmental damage and stress, maintains homeostasis, and provides key immune functions [1]. The skin consists of three main layers: the epidermis, which is the outermost protective layer composed of corneocytes, keratinocytes, melanocytes, and Langerhan’s cells; the dermis, which imparts the elasticity and structure of the skin by the extracellular matrix proteins such as collagen and elastin secreted by fibroblasts; and subcutaneous tissue, which is composed mostly of adipose tissue, containing adipocytes and stromal vascular cells [2]. Over time, skin undergoes aging due to intrinsic factors resulting in cell senescence characterized by reduced collagen production and decreased elasticity, as well as due to extrinsic factors like UV exposure, pollution, and oxidative stress [2].

To combat the effects of skin aging, modern skincare aims to restore skin hydration and elasticity, reduce pigmentation, and prevent or repair damage caused by aging and environmental factors. According to McKinsey, the global cosmetics market is a multi-billion industry which grew to $446 billion in 2023 and is estimated to reach $590 billion by 2028. It is characterized by a CAGR of 6%, with skincare representing the largest share of cosmetics, accounting for 44% of the market in 2023 [3]. There has been renewed interest in natural products for cosmetics, both from consumers and industry due to potential risks associated with synthetic compounds, as well as the purported benefits of natural products [4].

Medicinal plants have been used in many cultures since ancient times and contain multifaceted therapeutic properties, which can be attributed to a multitude of secondary metabolite phytochemicals [4]. *Centella asiatica*, also known as Cica, is commonly used in traditional herbal medicine such as Chinese traditional medicine and Indian Ayurveda therapy. The plant contains a rich assortment of phytochemicals such as triterpenoids, flavonoids, and polyphenols that contribute to its anti-inflammatory, wound-healing, and central nervous system protective properties [5-7]. Due to its broad spectrum benefits, commercial products containing Cica extracts are available for wound healing and skin care purposes [8]. A cursory glance at major cosmetics retailers in the United States and Japan revealed a wide variety of Cica-based skin care products, representing a sizeable section of natural skin care market [9, 10].

Extracellular vesicles (EVs) are nanoscale lipid membrane-bound particles released by cells into the extracellular environment, carrying bioactive molecules that can modulate the physiological processes of target cells, even across disparate kingdoms of life [11]. Recent studies have shown that exosome-like EVs from plants, including Cica, promote skin improvement and wound healing more effectively than conventional plant extracts [12-14]. We have previously characterized ExoBella^®^, a proprietary preparation of EVs isolated from tissue culture Cica, EVs and confirmed their potent *in-vitro* antioxidant, anti-melanogenic, and anti-inflammatory properties [15]. Kristen Claire Supreme Rejuvenation Essence^®^ is a skincare product formulated with ExoBella^®^ Cica EVs as the main active ingredient. This clinical study aimed to evaluate skin improvement in volunteer users of Kristen Claire Supreme Rejuvenation Essence^®^ over a period of 28 days.

## Materials and Methods

### Participants and Study Design

Participants were healthy adults of any gender from the age of 18 to 60 years (inclusive). The inclusion criteria included the absence of chronic diseases, major illnesses, or allergies, and not currently using any medications or other skincare products. The participants were required to conduct a pre-test screening to confirm the absence of skin irritation or allergic reaction to the test product which consisted of applying a skin patch infused with the test product for 24 hours on the inner forearms.

After screening, participants were asked to use the test product twice daily (morning and evening) after cleansing their face without using exfoliating products for 28 consecutive days. Each application involved 2 drops, evenly applied to the face using fingertips for absorption. Product usage was self-recorded, and the products were stored at room temperature.

The skin quality tests were conducted at the investigation site Hungkuang University (Taichung City, Taiwan) onsite by the principal investigator. Each participant was to allocate 1 hour per test (including cleansing and waiting time). Skin quality tests were performed as a baseline test on day 0 (before using the test product), and subsequent tests at 7, 14, 21, and 28 days after test product use.

Measurements of the skin quality tests included skin hydration, melanin content, and elasticity index using a C+K Multi Probe Adaptor MPA580 skin analysis device (Courage-Khazaka Electronic GmbH, Koln, Germany) equipped with probes for hydration, pigmentation, and elasticity measurements. Probes were placed 1 cm horizontally and 1 cm vertically from the outer corner of each eye (on the left and right cheeks) and 2 cm above the midpoint of the eyebrows (forehead). Additionally, the VISIA^®^ Skin Analysis system (Canfield Imaging Systems, Fairfield, NJ, USA) was used to assess changes in wrinkle percentage, redness area percentage, and pore percentage by scanning full-face frontal and 45° angles on both sides of the face.

The tests were conducted in a controlled environment at 20 ± 1°C and 50 ± 5% relative humidity, without direct sunlight or air conditioning airflow. All measurements were non-invasive. Probes contact the skin’s surface to assess parameters based on electrical resistance, optical principles, and physical changes. Participants’ test results and photographic data were collected. This clinical study was registered at clinicaltrial.gov: NCT06850935

### Description of Test Product

The test product (Kristen Claire Supreme Rejuvenation Essence^®^) is a proprietary water-based formulation containing the main active ingredient ExoBella^®^, a preparation of EVs derived from tissue culture *C. asiatica*. Other minor active ingredients include *Avena sativa* kernel extract, *Chamomilla recutita* flower extract, *Epilobium angustifolium* flower/leaf/stem extract, allantoin, ectoin, sodium hyaluronate, and arginine. The appearance is clear with slightly gel-like in consistency with faint fragrance.

### Assessment of Outcomes

Skin hydration was measured using the Corneometer^®^ CM825 (Courage-Khazaka Electronic GmbH, Koln, Germany), which evaluates the dielectric constant related to epidermal moisture at a depth of 60–100 μm. Mexameter^®^ MX18 (Courage-Khazaka Electronic GmbH, Koln, Germany) was used to measure melanin content, which utilizes RGB light absorption (568 nm, 660 nm, and 880 nm) to measure melanin and hemoglobin content. Elasticity index was measured using the Cutometer^®^ Dual MPA580 system (Courage-Khazaka Electronic GmbH, Koln, Germany), based on suction and stretching properties of the skin.

VISIA^®^ Skin Analysis System (Canfield Imaging Systems, Fairfield, NJ, USA) was used to evaluate the following parameters:

- Wrinkle percentage was measured by using standard white light to detect shadow variations to determine the distribution and number of wrinkles. Fewer wrinkles yield higher percentages.
- Redness percentage was measured by RBX polarized light to identify vascular or inflammatory issues. Smaller red zones result in higher percentages.
- Pore percentage was measured under white light to analyze shadowed pore depressions and darker areas relative to surrounding skin. Fewer pores yield higher percentages.

### Data Presentation and Statistical Analysis

For each of the measured parameters, results are converted to skin improvement rate (%) calculated as follows:

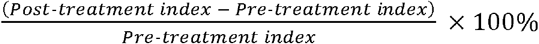

Where the pre-treatment index is the score of the parameter examined on Day 0, and the post-treatment index is the score of the parameter examined on Day 7, 14, 21, or 28. In skin hydration and elasticity, a positive skin improvement rate indicates higher hydration and high elasticity, respectively, when compared to Day 0. In skin melanin content, the skin improvement rate is negative because the parameter measures the decrease in melanin content and the lower skin improvement rate indicates greater decrease in melanin. In parameters measured using the VISIA^®^ Skin Analysis System, a higher skin improvement rate indicates decreased wrinkles, redness, or pores.

Results are presented as mean of the skin improvement rate with 95% confidence intervals shown as error bars. The analysis package in GraphPad Prism 6.01 (Boston, MA) was used for statistical analysis. A paired Student’s t-test was used to compare the difference between means of skin improvement rate on Days 7, 14, 21, or 28 against Day 0 (baseline).

## Results

### Participants

From September 4, 2024, to October 4, 2024, 20 healthy participants (4 males and 16 females; average age 36.5) were enrolled and all were assessed negative for irritation or allergic reactions to the test product prior to the study. The participants ranged from the age of 23 to 54 years, and the mean age was 36.5 with a standard deviation of 9.2.

### Assessments of Participants

The participants were assessed for changes in skin parameters, including skin hydration, melanin content, skin elasticity, wrinkle, redness (erythema), and skin pores. The raw data values of the assessment are presented in Tables S1 to S6. The evaluation was performed on three facial regions (left cheek, right cheek, and forehead) every 7 days over a 28-day period. In skin hydration, statistical significant increases in hydration percentage were observed across all facial regions (left cheek, right cheek, and forehead) over the course of 28 days compared to baseline, with the right cheek having slightly less improvement compared to other regions (Figure 1). Skin hydration improved by 3.1% to 5.1%, 5.8% to 10.6%, 9.6% to 15.2%, and 14.6% to 21.2% on Days 7, 14, 21, and 28, respectively.

**Figure 1.**
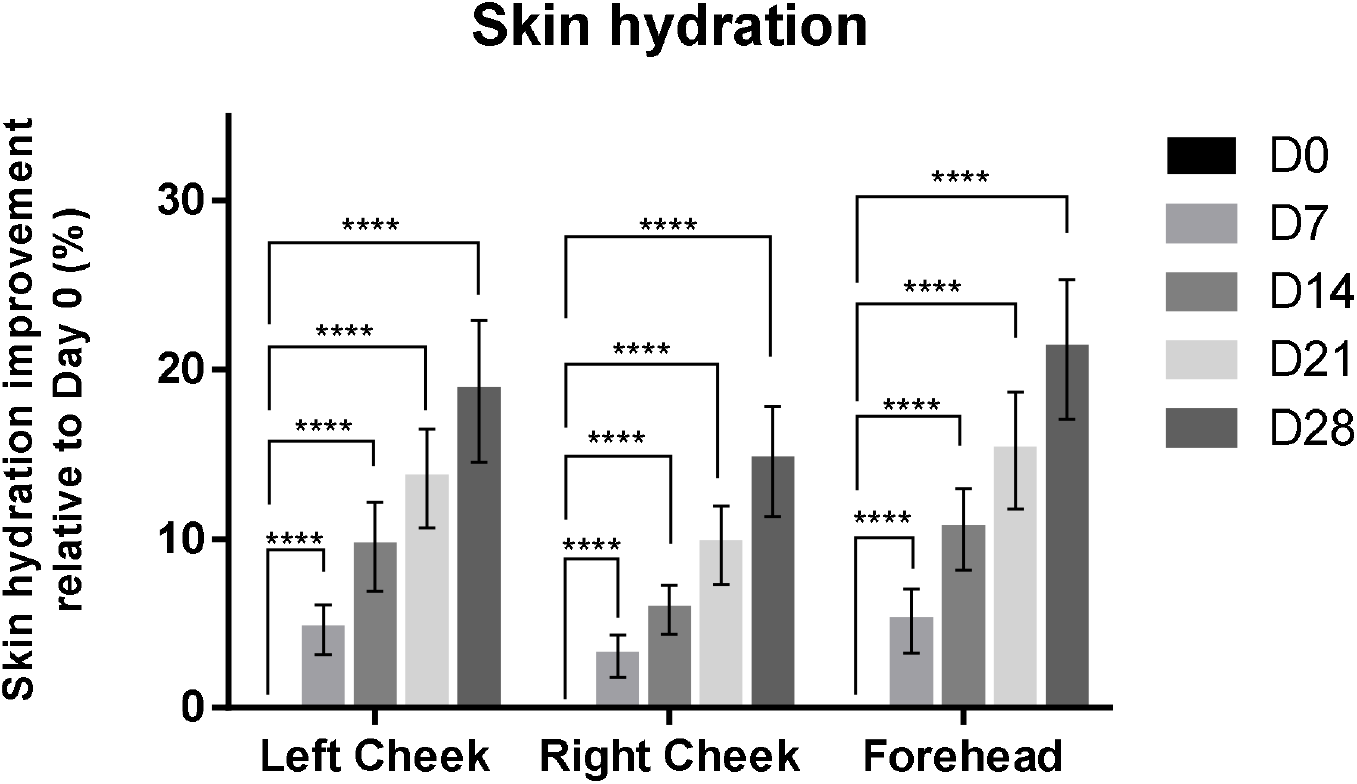
Change in skin hydration at 0, 7, 14, 21, and 28 days after treatment of test product. Higher values indicate more hydrated skin. Results are shown as bars representing means with error bars representing 95% confidence interval of the mean. Statistical significances on days 7, 14, 21, and 28 were calculated by paired t-test compared to value on day 0 (baseline). * P<0.05, ** P<0.01, *** P<0.001, *** P<0.0001

The treatment also resulted in significantly lowered melanin content throughout the study period with melanin decreasing by 1.2% to 2.3%, 4.1% to 5.0%, 6.2% to 7.8%, and 9.2% to 11.3%, on Days 7, 14, 21, and 28, respectively (Figure 2).

**Figure 2.**
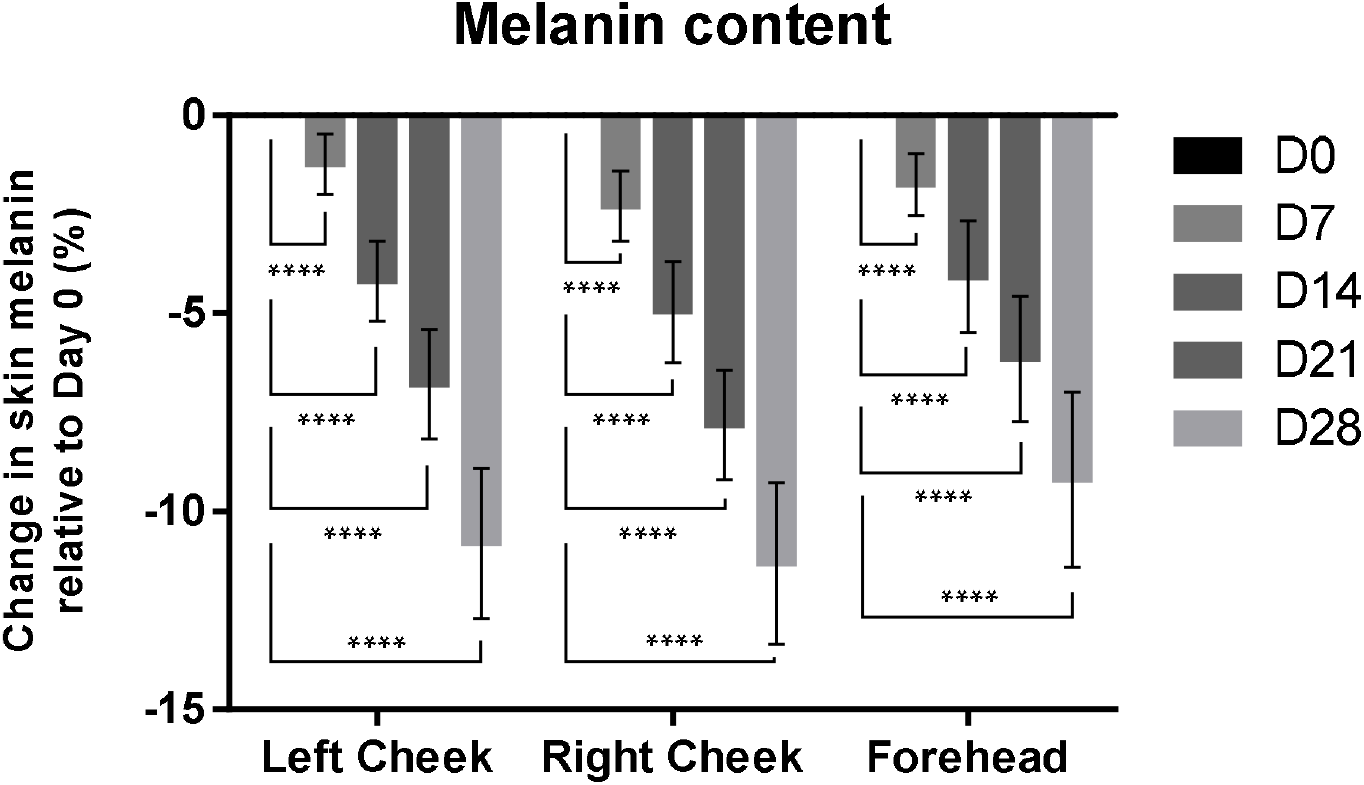
Change in skin melanin content at 0, 7, 14, 21, and 28 days after treatment of test product. Lower values indicate decreasing melanin content. Results are shown as bars representing means with error bars representing 95% confidence interval of the mean. Statistical significances on days 7, 14, 21, and 28 were calculated by paired t-test compared to value on day 0 (baseline). * P<0.05, ** P<0.01, *** P<0.001, *** P<0.0001’

Similarly, treatment resulted in significant improvement in skin elasticity over time uniformly across all regions tested (Figure 3). The improvement in skin elasticity was 2.2% to 3.5%, 4.6% to 5.9%, 7.4% to 8.9%, and 12.0% to 12.5% on Days 7, 14, 21, and 28, respectively.

**Figure 3.**
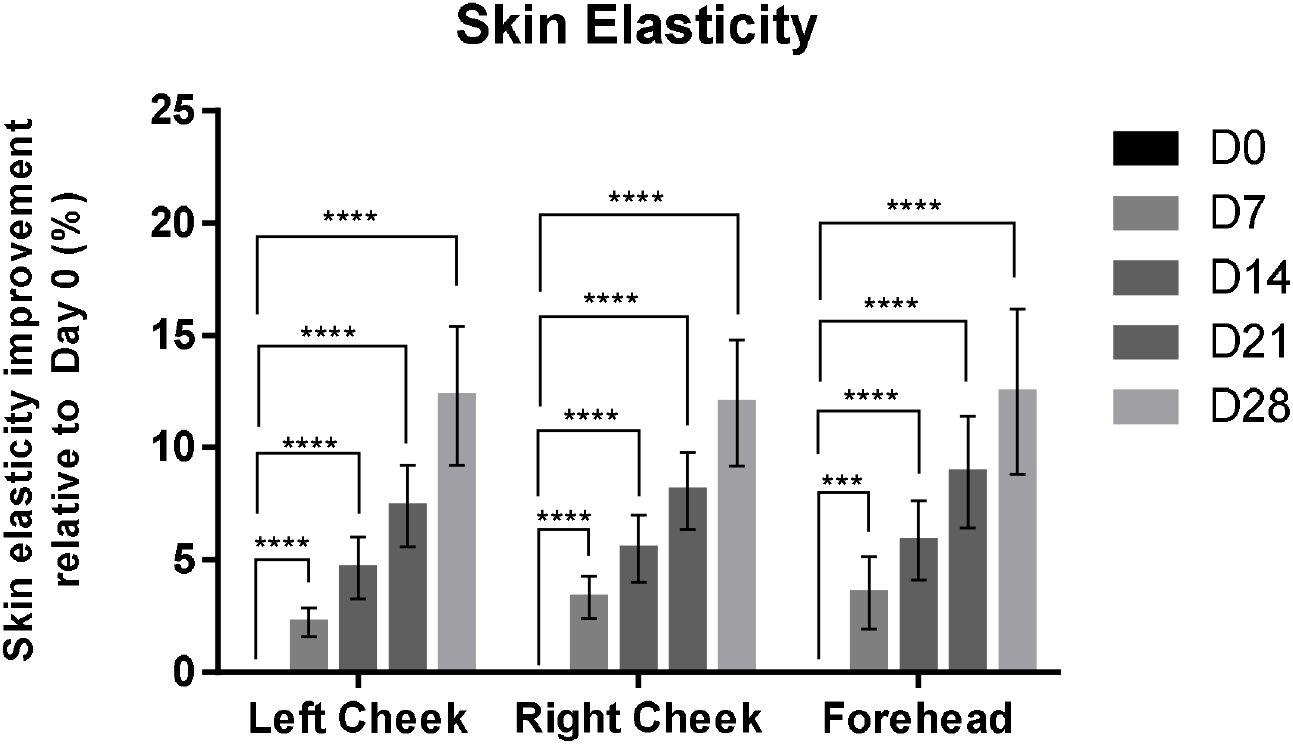
Change in skin elasticity at 0, 7, 14, 21, and 28 days after treatment of test product. Higher values indicate increasing skin elasticity. Results are shown as bars representing means with error bars representing 95% confidence interval of the mean. Statistical significances on days 7, 14, 21, and 28 were calculated by paired t-test compared to value on day 0 (baseline). * P<0.05, ** P<0.01, *** P<0.001, *** P<0.0001

The VISIA^®^ Skin Analysis System was used to image and analyze skin wrinkles, redness, and pore distribution simultaneously. Skin wrinkle condition improved by 8.6% to 16.9%, 19.3% to 24.9%, 28.2% to 30.0%, and 32.9% to 34.8% on Days 7, 14, 21, and 28, respectively (Figure 4). The forehead region lagged behind in wrinkle improvement at an earlier stage during the study but increased to approximately equal to that of other facial regions by Day 21 (Figure 4).

**Figure 4.**
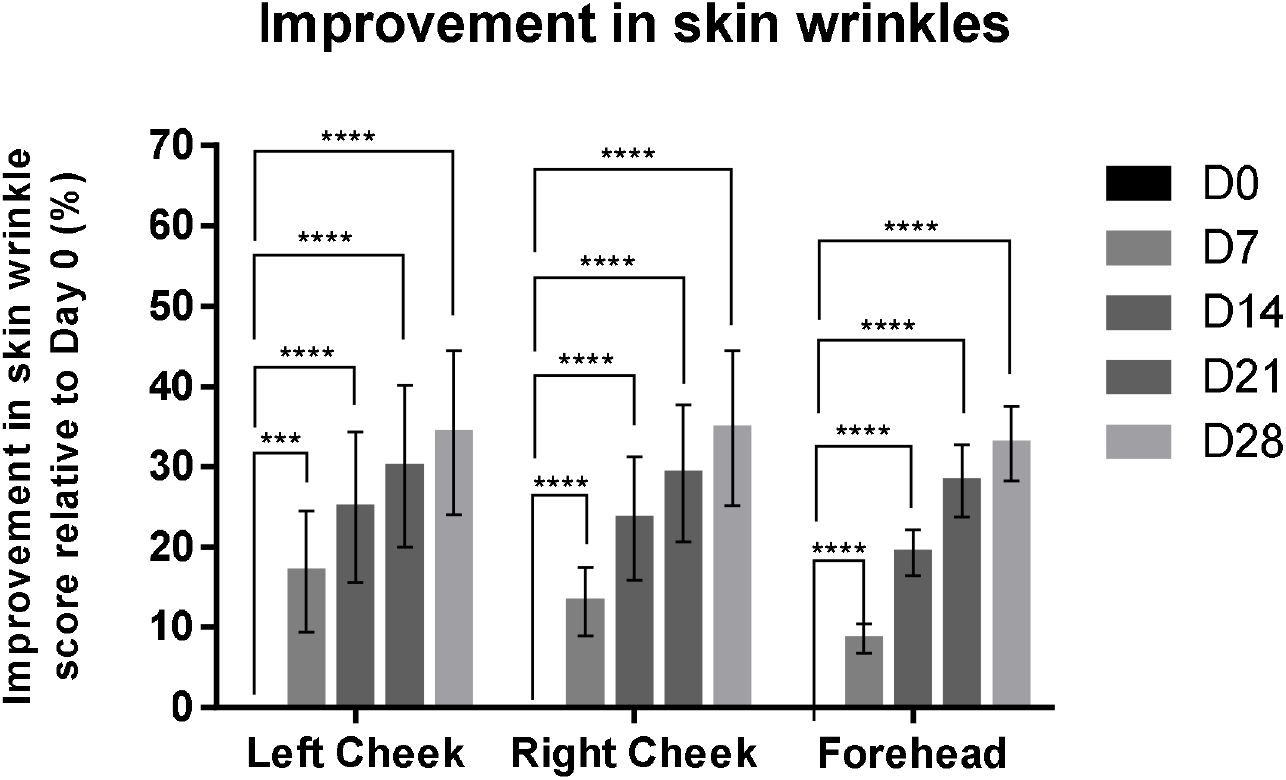
Change in skin wrinkles at 0, 7, 14, 21, and 28 days after treatment of test product. Higher values indicate decreasing distribution and number of wrinkles. Results are shown as bars representing means with error bars representing 95% confidence interval of the mean. Statistical significances on days 7, 14, 21, and 28 were calculated by paired t-test compared to value on day 0 (baseline). * P<0.05, ** P<0.01, *** P<0.001, *** P<0.0001

Skin redness was ameliorated during the study period by 5.6% to 7.9%, 13.1% to 14.6%, 19.9% to 22.7%, and 26.3% to 34.0% on Days 7, 14, 21, and 28, respectively (Figure 5).

**Figure 5.**
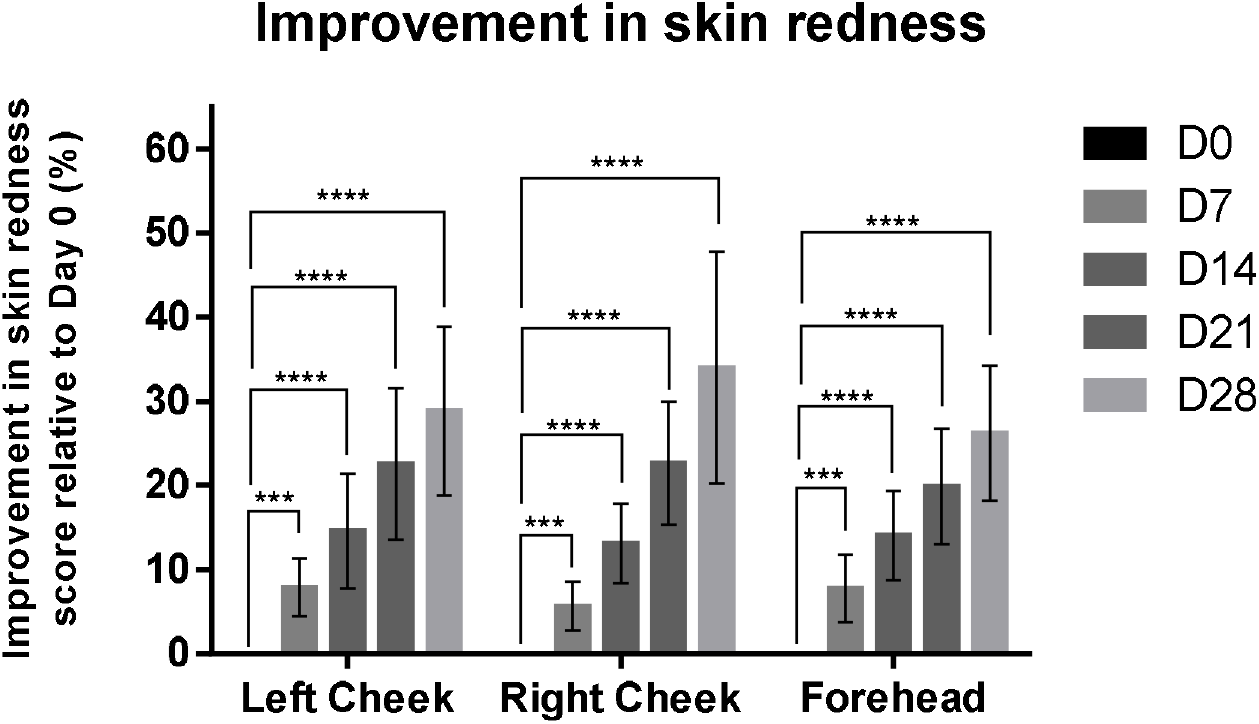
Change in skin redness at 0, 7, 14, 21, and 28 days after treatment of test product. Higher values indicate decreasing zone of redness. Results are shown as bars representing means with error bars representing 95% confidence interval of the mean. Statistical significances on days 7, 14, 21, and 28 were calculated by paired t-test compared to value on day 0 (baseline). * P<0.05, ** P<0.01, *** P<0.001, *** P<0.0001

Finally, skin pores reduction by treatment resulted in skin improvement by 9.3% to 12.4%, 19.8% to 21.8%, 28.5% to 32.1%, and 40.6% to 41.3% on Days 7, 14, 21, and 28, respectively (Figure 6). In summary, the treatment had a significant improvement in skin hydration and elasticity, while reducing the distribution and area of melanin, wrinkles, redness, and pores.

**Figure 6.**
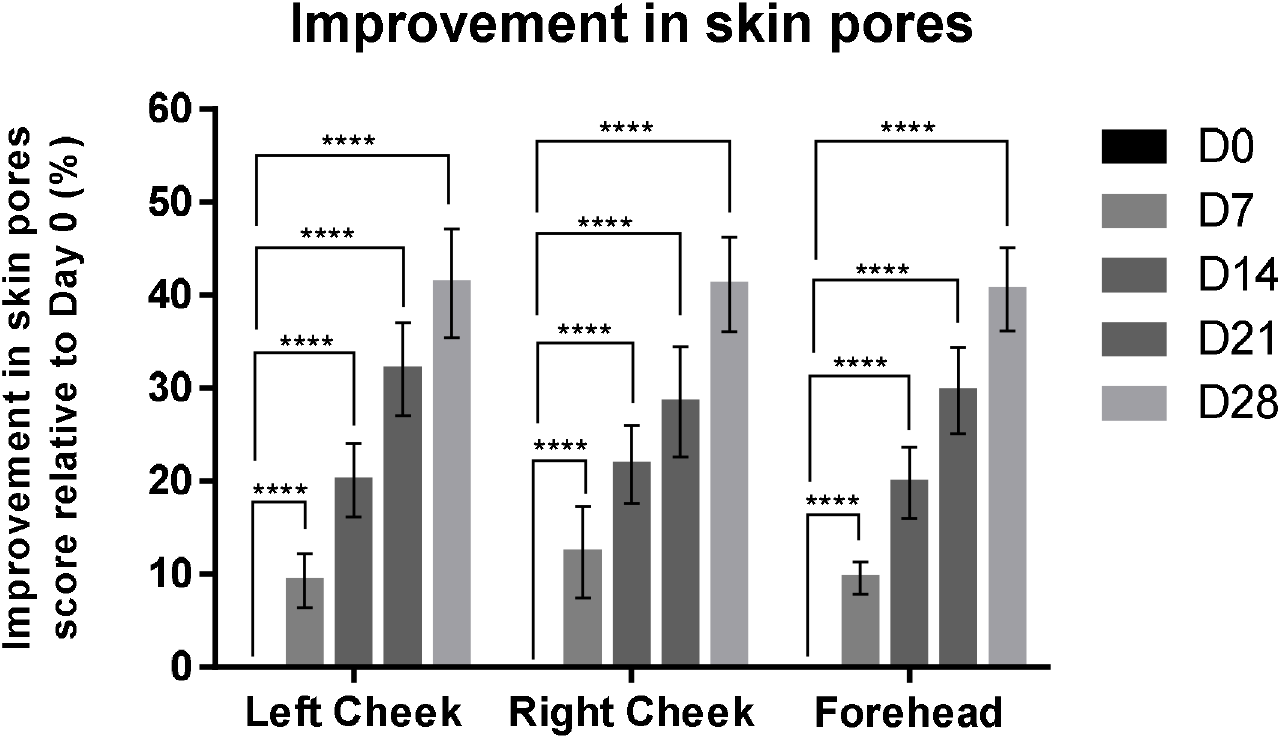
Change in skin pores at 0, 7, 14, 21, and 28 days after treatment of test product. Higher values indicate decreasing number and area of pores. Results are shown as bars representing means with error bars representing 95% confidence interval of the mean. Statistical significances on days 7, 14, 21, and 28 were calculated by paired t-test compared to value on day 0 (baseline). * P<0.05, ** P<0.01, *** P<0.001, *** P<0.0001

To investigate the effect of treatment in different age groups, the data was stratified according to the age of participants into two groups: > 40 years and < 40 years. The means of skin improvement rate of each parameter were compared between the two age groups at the three facial regions. We found no significant differences in skin hydration and elasticity between the two age groups (Figure S1). However, those aged above 40 years had significant less reduction in melanin in both cheek regions compared to participants aged less than 40 years on Day 28 of the study, while there were no significant differences between forehead region in both age groups (Figure S1). For parameters analyzed with VISIA^®^ System, there were no significant differences between the two age groups in changes in skin wrinkles and pores (Figure S2). In redness skin area, the overall improvement in > 40 years age group was less than that of < 40 years age group, and the difference between two age groups was significant on Day 28 at the forehead region (Figure S2).

## Discussion

In this small-scale clinical study, we observed positive improvements on the skin in participants treated with Cica EV-based test product. Skin integrity parameters such as hydration and elasticity increased after treatment with the test product. According to our previous study, Cica EVs could achieve improvement in skin hydration via upregulation of water retention genes such as aquaporin and filaggrin, and higher skin elasticity by increasing production of collagen and elastin [15]. The triterpene madecassoside in Cica has already been studied for its anti-melanogenic effect via anti-inflammatory effects [16]. Besides madecassoside, miRNAs within Cica EVs can also target melanogenic genes such as tyrosinase and other pathway inducers (WNT, CRTC1), as has been observed in our previous study and others [13, 15].

Cica extracts have already been used in commercial skin care products due to their skin healing, antioxidant, and anti-melanogenic properties imparted by their bioactive compounds [6-8]. The mechanisms through which exosomes promote skin regeneration include angiogenesis, collagen synthesis, and regulation of inflammation [17, 18]. A study discovered 11 novel miRNAs contained within Cica exosomes, which were predicated to inhibit genes involved in melanin production and dermatitis [13]. The study also found that Cica exosomes induced higher gene expression than *C. asiatica* extracts, possibly due to better penetration of skin cells by exosomes and the presence of packaged miRNAs [13]. In formulation of the test product, ExoBella^®^ Cica EVs may work in concert with other herbal extracts and small molecule compounds (arbutin, niacinamide, vitamin E) to achieve the results seen in this study.

The main limitation of the study is the small sample size, which may not include sufficient diversity of demographics such as different skin types and ethnicities due to the homogeneity of population in Taiwan. The small sample size also made it unfeasible to conduct a more robust analysis of differences between age groups, and stratification into smaller age groups (e.g. 16∼ 25 years, 26∼35 years, etc.). The sample population is also heavily gender-skewed toward female (80% of participants), which can also influence the generalizability of the results. The other limitation was the relatively short duration of the study (28 days). This may not be sufficient to capture the long-term effects of the test product as skin quality can be influenced by numerous factors over time including aging, UV exposure, and other environmental factors. A longer study period and follow-up would allow a more comprehensive understanding of the product’s sustained efficacy and potential adverse effects. Lastly, as this was not a placebo-controlled nor double-blinded study, it would be difficult to determine whether the improvements observed were attributed to the use of test product, natural improvement of skin over time, or due to other external factors.

In conclusion, the results of this study support the positive effects on skin quality of Kristen Claire Supreme Rejuvenation Essence^®^ in improving skin hydration and elasticity while reducing melanin, wrinkles, redness, and pores. The differences observed between age groups and the three facial regions were minimal. Future studies should address the above mentioned limitations to provide clearer insights into the long-term benefits on skin improvement and application of the test product across more diverse set of population to address a global market.

## Supplementary materials

Figure S1: Change in skin hydration, melanin, and elasticity at 7, 14, 21, and 28 days after treatment of test product in left cheek, right cheek, and forehead facial regions grouped by age (< 40 years vs. > 40 years).

Figure S2: Change in skin wrinkles, redness, and pores at 7, 14, 21, and 28 days after treatment of test product in left cheek, right cheek, and forehead facial regions grouped by age (< 40 years vs. > 40 years).

Table S1: Data values for skin hydration during the study period

Table S2: Data values for skin melanin content during the study period.

Table S3: Data values for skin elasticity during the study period.

Table S4: Wrinkle score during the study period.

Table S5: Redness score during the study period.

Table S6: Pore score during the study period.

## Author contributions

Conceptualization, supervision and project administration: T.-M.C., C.C., T.-Y.K.

Visualization and formal analysis: T.-M.C., L.T.-C.L.

Investigation, data acquisition and curation: T.-M.C., H.-C.H., S.-S.W.

Methodology: T.-M.C., C.-C.W., H.-C.H., S.-S.W., C.-H.C., P.-L.K., W.-H.T., W.-Y.Q. I.P., T.-Y.K.

Resources: C.-C.W., C.-H.C., P.-L.K., W.-H.T., T.-Y.K.

Writing—original draft preparation: T.-.M.C., L.T.-C.L.

Writing—review and editing: T.-M.C., L.T.-C.L., W.-Y.Q., I.P., C.C., T.-Y.K.

All authors reviewed the manuscript

## Funding

Schweitzer Biotech Company provided the necessary funding for the study as well as provided test product Kristen Claire Supreme Rejuvenation Essence^®^, but had otherwise no role in enrollment, participation, data collection, data analysis, or completion of final report of the clinical study.

## Institutional Review Board Statement

The final protocol, case report form, and informed consent form were reviewed and approved by the institutional review board of the Antai Tian-Sheng Memorial Hospital (IRB No. 24-064-A, approved May 16, 2024). All participants provided written informed consent prior to the start of the study. This study was conducted in compliance with Declaration of Helsinki and in compliance with all International Conference on Harmonization Good Clinical Practice Guidelines (ICH-GCP).

## Informed Consent Statement

All participants provided written informed consent prior to the start of the study.

## Data Availability

All data generated or analyzed during this study are included in this manuscript.

## Acknowledgements

We thank Dr. Meei-Yun Lin of Schweitzer Biotech Company for reading and providing feedback for preparation of the manuscript.

## Conflicts of Interest

C.-C. W., C.-H. C., P.-L. K., W.-H. T., L. T.-C. L., C. C., and T.-Y. K. are employees of Schweitzer Biotech Company. T.-M. C. and S.-S. W. have no competing interests to declare. This study and the test product were funded and provided by Schweitzer Biotech Company.

## Notes

### Clinical Trial

NCT06850935

### Author Declarations

The final protocol, case report form, and informed consent form were reviewed and approved by the institutional review board of the Antai Tian-Sheng Memorial Hospital (IRB No. 24-064-A, approved May 16, 2024).

### Summary of Updates

Correction to test product ingredient list.

