## Supplementary Tables for "Evaluation of Effects on Skin Quality of a *Centella asiatica* Extracellular Vesicle-based Skin Care Formulation: A 28-Day Facial Skin Quality Study"

Table S1. Data values (a.u.) for skin hydration during the study period.

| **Left cheek** | | | | | |
| --- | --- | --- | --- | --- | --- |
| **Participant #** | **Day 0** | **Day 7** | **Day 14** | **Day 21** | **Day 28** |
| 1 | 64.7 | 65.9 | 71.4 | 71.9 | 72.2 |
| 2 | 65.9 | 67.2 | 67.9 | 69.5 | 72.1 |
| 3 | 66.6 | 70.3 | 72.6 | 74.5 | 76.7 |
| 4 | 71.3 | 74.8 | 75.7 | 77.8 | 79.3 |
| 5 | 75.9 | 79.3 | 80.1 | 81.5 | 90.6 |
| 6 | 71.5 | 73.2 | 76 | 78.1 | 78.4 |
| 7 | 72.8 | 73.2 | 74.9 | 80.2 | 83.4 |
| 8 | 72.9 | 73 | 73.5 | 75.7 | 75.8 |
| 9 | 54.5 | 55.8 | 60.7 | 66.8 | 74.9 |
| 10 | 74.9 | 78.6 | 78.9 | 81.5 | 83.1 |
| 11 | 61.4 | 65.3 | 70.2 | 74.4 | 78.9 |
| 12 | 63.2 | 69.9 | 77.6 | 79.1 | 82.9 |
| 13 | 71.3 | 78 | 80.9 | 81.3 | 82.2 |
| 14 | 71.8 | 75.9 | 78.4 | 80.2 | 80.3 |
| 15 | 69.6 | 70.3 | 71.2 | 75.3 | 83.7 |
| 16 | 57.6 | 61.6 | 64.5 | 67.3 | 73.4 |
| 17 | 60.3 | 60.6 | 70.9 | 71.3 | 72.6 |
| 18 | 59.8 | 65.3 | 65.5 | 68.2 | 73.2 |
| 19 | 60.1 | 63.6 | 68.6 | 72.5 | 79 |
| 20 | 60.7 | 65.3 | 69.5 | 73.8 | 74.3 |
| Mean (SD) | 66.3 (6.36) | 69.4 (6.47) | 72.5 (5.47) | 75.0 (4.84) | 78.4 (4.95) |
| **Right cheek** | | | | | |
| **Participant #** | **Day 0** | **Day 7** | **Day 14** | **Day 21** | **Day 28** |
| 1 | 64.3 | 65.3 | 66.7 | 71 | 74 |
| 2 | 59.7 | 60.3 | 62.3 | 70.2 | 71.3 |
| 3 | 70.1 | 70.8 | 76 | 76.7 | 78.8 |
| 4 | 77.8 | 78.5 | 79.3 | 80.1 | 80.4 |
| 5 | 77.4 | 78.4 | 82.6 | 83.4 | 88.4 |
| 6 | 73.6 | 73.7 | 74.8 | 76.6 | 77.8 |
| 7 | 76.1 | 77.1 | 80.3 | 80.5 | 82.5 |
| 8 | 70.3 | 71.5 | 72.3 | 73.7 | 80.1 |
| 9 | 60.1 | 62.8 | 63.6 | 70.6 | 74.8 |
| 10 | 74.6 | 76.7 | 77.5 | 78.2 | 79.2 |
| 11 | 63.3 | 64.9 | 65.7 | 74.8 | 75.9 |
| 12 | 74.9 | 75.5 | 78.2 | 80.6 | 83.9 |
| 13 | 73.4 | 76.1 | 77.9 | 78.1 | 82.6 |
| 14 | 79.6 | 80.2 | 81.3 | 81.7 | 83.1 |
| 15 | 71.5 | 73.6 | 74.6 | 76.9 | 82.6 |
| 16 | 62.9 | 65.7 | 68.3 | 71.7 | 78.7 |
| 17 | 64.6 | 70.5 | 71.6 | 72.5 | 73.2 |
| 18 | 57.8 | 61.9 | 64.8 | 65.3 | 73.2 |
| 19 | 62.2 | 65 | 66.6 | 69.7 | 76.7 |
| 20 | 64.5 | 70.3 | 71.9 | 73.4 | 74.7 |
| Mean (SD) | 68.9 (6.86) | 70.9 (6.18) | 72.8 (6.36) | 75.3 (4.77) | 78.6 (4.42) |
| **Forehead** | | | | | |
| **Participant #** | **Day 0** | **Day 7** | **Day 14** | **Day 21** | **Day 28** |
| 1 | 55.5 | 57.7 | 67.4 | 71.3 | 71.6 |
| 2 | 51.5 | 52.1 | 56.8 | 60.9 | 67.4 |
| 3 | 45.5 | 50.3 | 53.9 | 56.8 | 60.1 |
| 4 | 64.1 | 65.5 | 67.8 | 71.8 | 72.1 |
| 5 | 64.5 | 72.7 | 74.7 | 75 | 81.8 |
| 6 | 70.7 | 72.3 | 74.7 | 75.4 | 77.6 |
| 7 | 65.8 | 66.2 | 70.2 | 70.7 | 72.3 |
| 8 | 68.3 | 69.9 | 70.7 | 71.9 | 73.5 |
| 9 | 60.2 | 63.1 | 66.9 | 67.8 | 71.8 |
| 10 | 62.9 | 67.9 | 70.3 | 72 | 74.5 |
| 11 | 55 | 57.8 | 58.7 | 60.8 | 63.7 |
| 12 | 68.7 | 69.6 | 73.1 | 78.3 | 80.9 |
| 13 | 56.7 | 58.4 | 63.8 | 67.5 | 69.2 |
| 14 | 70.5 | 75.3 | 76.2 | 77.4 | 78.6 |
| 15 | 52.7 | 54.3 | 60.7 | 65.2 | 70.1 |
| 16 | 44.4 | 51.3 | 53 | 58.7 | 59.3 |
| 17 | 56.7 | 60.7 | 62.3 | 65.2 | 73.6 |
| 18 | 54.7 | 58.5 | 60.4 | 60.6 | 71.7 |
| 19 | 58.2 | 59.1 | 62.3 | 67.5 | 69.2 |
| 20 | 59.5 | 61.7 | 62.4 | 63.6 | 68 |
| Mean (SD) | 59.3 (7.67) | 62.2 (7.48) | 65.3 (6.95) | 67.9 (6.35) | 71.4 (5.99) |

Table S2. Data values for skin melanin content during the study period.

| **Left cheek** | | | | | |
| --- | --- | --- | --- | --- | --- |
| **Participant #** | **Day 0** | **Day 7** | **Day 14** | **Day 21** | **Day 28** |
| 1 | 150 | 149 | 145 | 140 | 134 |
| 2 | 176 | 173 | 168 | 165 | 155 |
| 3 | 250 | 249 | 240 | 232 | 224 |
| 4 | 242 | 240 | 231 | 225 | 216 |
| 5 | 149 | 146 | 140 | 132 | 135 |
| 6 | 158 | 157 | 151 | 147 | 142 |
| 7 | 160 | 158 | 152 | 150 | 141 |
| 8 | 153 | 152 | 150 | 146 | 140 |
| 9 | 149 | 145 | 141 | 138 | 132 |
| 10 | 189 | 187 | 182 | 179 | 173 |
| 11 | 171 | 170 | 165 | 162 | 156 |
| 12 | 118 | 115 | 113 | 110 | 103 |
| 13 | 145 | 144 | 140 | 136 | 124 |
| 14 | 165 | 164 | 160 | 155 | 147 |
| 15 | 167 | 165 | 161 | 156 | 151 |
| 16 | 173 | 171 | 167 | 161 | 155 |
| 17 | 165 | 164 | 157 | 154 | 149 |
| 18 | 107 | 105 | 101 | 98 | 90 |
| 19 | 141 | 140 | 136 | 132 | 128 |
| 20 | 147 | 143 | 139 | 136 | 131 |
| Mean (SD) | 163.8 (33.8) | 161.9 (34.1) | 157.0 (32.6) | 152.7 (31.8) | 146.3 (31.3) |
| **Right cheek** | | | | | |
| **Participant #** | **Day 0** | **Day 7** | **Day 14** | **Day 21** | **Day 28** |
| 1 | 157 | 154 | 152 | 146 | 139 |
| 2 | 174 | 170 | 167 | 161 | 153 |
| 3 | 235 | 232 | 229 | 221 | 216 |
| 4 | 235 | 230 | 226 | 222 | 217 |
| 5 | 148 | 145 | 142 | 136 | 132 |
| 6 | 159 | 156 | 150 | 148 | 141 |
| 7 | 154 | 147 | 145 | 140 | 134 |
| 8 | 151 | 148 | 143 | 140 | 135 |
| 9 | 154 | 149 | 145 | 141 | 136 |
| 10 | 187 | 183 | 180 | 174 | 165 |
| 11 | 170 | 168 | 163 | 159 | 152 |
| 12 | 124 | 121 | 116 | 112 | 104 |
| 13 | 147 | 144 | 140 | 134 | 129 |
| 14 | 162 | 158 | 152 | 146 | 143 |
| 15 | 164 | 160 | 156 | 151 | 146 |
| 16 | 172 | 165 | 160 | 156 | 153 |
| 17 | 166 | 162 | 155 | 150 | 143 |
| 18 | 112 | 109 | 105 | 102 | 97 |
| 19 | 145 | 142 | 140 | 137 | 134 |
| 20 | 145 | 144 | 137 | 134 | 130 |
| Mean (SD) | 163.1 (29.7) | 159.4 (29.4) | 155.2 (29.7) | 150.5 (29.0) | 150.0 (28.9) |
| **Forehead** | | | | | |
| **Participant #** | **Day 0** | **Day 7** | **Day 14** | **Day 21** | **Day 28** |
| 1 | 207 | 205 | 201 | 197 | 194 |
| 2 | 209 | 207 | 204 | 200 | 192 |
| 3 | 288 | 285 | 281 | 276 | 267 |
| 4 | 274 | 271 | 265 | 260 | 256 |
| 5 | 137 | 134 | 130 | 127 | 121 |
| 6 | 133 | 131 | 128 | 125 | 117 |
| 7 | 185 | 182 | 176 | 173 | 171 |
| 8 | 179 | 177 | 173 | 170 | 163 |
| 9 | 198 | 195 | 190 | 188 | 182 |
| 10 | 221 | 219 | 217 | 210 | 203 |
| 11 | 250 | 247 | 241 | 237 | 232 |
| 12 | 146 | 141 | 136 | 133 | 124 |
| 13 | 214 | 210 | 206 | 200 | 193 |
| 14 | 202 | 197 | 195 | 193 | 184 |
| 15 | 169 | 164 | 158 | 153 | 150 |
| 16 | 231 | 228 | 222 | 217 | 213 |
| 17 | 237 | 230 | 225 | 220 | 215 |
| 18 | 173 | 169 | 163 | 160 | 155 |
| 19 | 172 | 168 | 162 | 157 | 152 |
| 20 | 175 | 173 | 170 | 165 | 161 |
| Mean (SD) | 200.0 (42.3) | 196.7 (42.3) | 192.2 (42.2) | 188.1 (41.6) | 182.3 (41.6) |

Table S3. Data values for skin elasticity (R2) during the study period.

| **Left cheek** | | | | | |
| --- | --- | --- | --- | --- | --- |
| **Participant #** | **Day 0** | **Day 7** | **Day 14** | **Day 21** | **Day 28** |
| 1 | 0.5438 | 0.5539 | 0.5583 | 0.5743 | 0.5914 |
| 2 | 0.6067 | 0.6087 | 0.6148 | 0.6215 | 0.6445 |
| 3 | 0.5118 | 0.5298 | 0.5631 | 0.5831 | 0.6148 |
| 4 | 0.5753 | 0.6049 | 0.6139 | 0.6205 | 0.6348 |
| 5 | 0.6117 | 0.6205 | 0.6221 | 0.6371 | 0.6554 |
| 6 | 0.5037 | 0.5135 | 0.5158 | 0.5279 | 0.5812 |
| 7 | 0.6151 | 0.6197 | 0.6247 | 0.6279 | 0.6378 |
| 8 | 0.5602 | 0.5719 | 0.5985 | 0.6049 | 0.6291 |
| 9 | 0.5657 | 0.5821 | 0.6037 | 0.6158 | 0.6408 |
| 10 | 0.5714 | 0.5825 | 0.5903 | 0.6144 | 0.6321 |
| 11 | 0.5298 | 0.5354 | 0.5487 | 0.6032 | 0.6149 |
| 12 | 0.5632 | 0.5781 | 0.5855 | 0.5906 | 0.6293 |
| 13 | 0.5652 | 0.5742 | 0.5894 | 0.6076 | 0.6149 |
| 14 | 0.5138 | 0.5341 | 0.5771 | 0.5943 | 0.6842 |
| 15 | 0.5663 | 0.5848 | 0.5931 | 0.6076 | 0.6272 |
| 16 | 0.6045 | 0.6098 | 0.6073 | 0.6169 | 0.634 |
| 17 | 0.5161 | 0.5248 | 0.538 | 0.5629 | 0.6145 |
| 18 | 0.5455 | 0.5731 | 0.5845 | 0.6061 | 0.6252 |
| 19 | 0.6045 | 0.6143 | 0.6294 | 0.6372 | 0.6483 |
| 20 | 0.6008 | 0.6045 | 0.6298 | 0.6371 | 0.6717 |
| Mean (SD) | 0.5638 (0.036) | 0.5760 (0.034) | 0.5894 (0.032) | 0.6045 (0.027) | 0.6313 (0.024) |
| **Right cheek** | | | | | |
| **Participant #** | **Day 0** | **Day 7** | **Day 14** | **Day 21** | **Day 28** |
| 1 | 0.5776 | 0.5837 | 0.5931 | 0.6041 | 0.6218 |
| 2 | 0.5379 | 0.5448 | 0.5537 | 0.5659 | 0.5989 |
| 3 | 0.5076 | 0.5249 | 0.5678 | 0.5759 | 0.6137 |
| 4 | 0.5081 | 0.5349 | 0.5637 | 0.5716 | 0.6073 |
| 5 | 0.6017 | 0.6198 | 0.6216 | 0.6372 | 0.6439 |
| 6 | 0.4532 | 0.4671 | 0.4836 | 0.5162 | 0.5686 |
| 7 | 0.5803 | 0.6074 | 0.6087 | 0.6138 | 0.6224 |
| 8 | 0.6024 | 0.625 | 0.6239 | 0.6435 | 0.6329 |
| 9 | 0.6138 | 0.6184 | 0.6248 | 0.6317 | 0.6482 |
| 10 | 0.5762 | 0.5932 | 0.6018 | 0.6125 | 0.6247 |
| 11 | 0.6071 | 0.6072 | 0.6178 | 0.6283 | 0.6331 |
| 12 | 0.6181 | 0.6285 | 0.6386 | 0.6468 | 0.6635 |
| 13 | 0.5957 | 0.6014 | 0.6157 | 0.6231 | 0.6374 |
| 14 | 0.5448 | 0.5631 | 0.5846 | 0.6029 | 0.6143 |
| 15 | 0.5163 | 0.5385 | 0.5433 | 0.5506 | 0.5841 |
| 16 | 0.5619 | 0.5881 | 0.5732 | 0.6019 | 0.6285 |
| 17 | 0.5026 | 0.5432 | 0.5513 | 0.5574 | 0.5928 |
| 18 | 0.5429 | 0.5654 | 0.5831 | 0.6076 | 0.6158 |
| 19 | 0.5416 | 0.5634 | 0.5793 | 0.6035 | 0.6232 |
| 20 | 0.5328 | 0.5667 | 0.5862 | 0.6017 | 0.6316 |
| Mean (SD) | 0.5561 (0.045) | 0.5742 (0.041) | 0.5858 (0.036) | 0.5998 (0.034) | 0.6203 (0.022) |
| **Forehead** | | | | | |
| **Participant #** | **Day 0** | **Day 7** | **Day 14** | **Day 21** | **Day 28** |
| 1 | 0.5734 | 0.5832 | 0.5914 | 0.6076 | 0.6184 |
| 2 | 0.5989 | 0.6018 | 0.6138 | 0.6216 | 0.6352 |
| 3 | 0.5069 | 0.5371 | 0.5544 | 0.5634 | 0.5931 |
| 4 | 0.6324 | 0.6512 | 0.6538 | 0.6578 | 0.6612 |
| 5 | 0.5643 | 0.5714 | 0.6051 | 0.6171 | 0.6296 |
| 6 | 0.5109 | 0.5701 | 0.5819 | 0.6016 | 0.6142 |
| 7 | 0.5633 | 0.5679 | 0.5731 | 0.5869 | 0.6065 |
| 8 | 0.5827 | 0.5941 | 0.5966 | 0.6173 | 0.6237 |
| 9 | 0.5809 | 0.6017 | 0.6147 | 0.6227 | 0.6356 |
| 10 | 0.6019 | 0.6127 | 0.6213 | 0.6331 | 0.6461 |
| 11 | 0.5839 | 0.5928 | 0.5991 | 0.6072 | 0.6128 |
| 12 | 0.5171 | 0.5232 | 0.5466 | 0.5525 | 0.6039 |
| 13 | 0.5424 | 0.5738 | 0.5843 | 0.6059 | 0.6132 |
| 14 | 0.4968 | 0.5148 | 0.5239 | 0.5674 | 0.5948 |
| 15 | 0.5381 | 0.5548 | 0.5748 | 0.5826 | 0.5913 |
| 16 | 0.5664 | 0.5703 | 0.5818 | 0.6077 | 0.6118 |
| 17 | 0.4632 | 0.5183 | 0.5234 | 0.5536 | 0.6083 |
| 18 | 0.4649 | 0.5027 | 0.5249 | 0.5611 | 0.6086 |
| 19 | 0.5872 | 0.5931 | 0.6052 | 0.6144 | 0.6296 |
| 20 | 0.5481 | 0.5581 | 0.5736 | 0.5826 | 0.5991 |
| Mean (SD) | 0.5512 (0.046) | 0.5697 (0.037) | 0.5822 (0.035) | 0.5982 (0.029) | 0.6169 (0.018) |

Table S4. Wrinkle score (%, higher = less wrinkles) during the study period.

| **Left cheek** | | | | | |
| --- | --- | --- | --- | --- | --- |
| **Participant #** | **Day 0** | **Day 7** | **Day 14** | **Day 21** | **Day 28** |
| 1 | 78 | 82 | 84 | 89 | 92 |
| 2 | 80 | 91 | 92 | 94 | 95 |
| 3 | 77 | 84 | 88 | 93 | 95 |
| 4 | 68 | 81 | 90 | 91 | 92 |
| 5 | 62 | 75 | 86 | 88 | 98 |
| 6 | 78 | 82 | 92 | 94 | 95 |
| 7 | 63 | 81 | 88 | 90 | 91 |
| 8 | 85 | 88 | 91 | 94 | 98 |
| 9 | 76 | 79 | 84 | 92 | 96 |
| 10 | 84 | 87 | 88 | 95 | 96 |
| 11 | 89 | 91 | 94 | 94 | 96 |
| 12 | 50 | 78 | 89 | 93 | 94 |
| 13 | 70 | 83 | 86 | 87 | 88 |
| 14 | 84 | 87 | 90 | 93 | 95 |
| 15 | 60 | 89 | 93 | 93 | 98 |
| 16 | 54 | 67 | 79 | 88 | 89 |
| 17 | 76 | 80 | 84 | 84 | 90 |
| 18 | 64 | 91 | 93 | 95 | 96 |
| 19 | 78 | 82 | 86 | 89 | 98 |
| 20 | 64 | 77 | 83 | 94 | 97 |
| Mean (SD) | 72.0 (10.90) | 82.8 (6.08) | 88.0 (3.99) | 91.5 (3.09) | 94.5 (3.10) |
| **Right cheek** | | | | | |
| **Participant #** | **Day 0** | **Day 7** | **Day 14** | **Day 21** | **Day 28** |
| 1 | 73 | 77 | 84 | 88 | 97 |
| 2 | 82 | 88 | 90 | 92 | 94 |
| 3 | 76 | 82 | 87 | 88 | 82 |
| 4 | 62 | 77 | 80 | 94 | 98 |
| 5 | 64 | 74 | 80 | 85 | 89 |
| 6 | 74 | 82 | 91 | 92 | 98 |
| 7 | 70 | 88 | 89 | 96 | 98 |
| 8 | 82 | 92 | 94 | 96 | 98 |
| 9 | 75 | 80 | 91 | 91 | 93 |
| 10 | 78 | 82 | 85 | 88 | 92 |
| 11 | 78 | 83 | 87 | 90 | 93 |
| 12 | 58 | 69 | 77 | 87 | 98 |
| 13 | 68 | 75 | 86 | 88 | 95 |
| 14 | 82 | 85 | 89 | 91 | 93 |
| 15 | 64 | 69 | 78 | 83 | 90 |
| 16 | 50 | 71 | 93 | 95 | 98 |
| 17 | 74 | 84 | 90 | 92 | 97 |
| 18 | 68 | 76 | 82 | 87 | 92 |
| 19 | 74 | 83 | 92 | 94 | 97 |
| 20 | 67 | 78 | 86 | 89 | 90 |
| Mean (SD) | 71.0 (8.45) | 79.8 (6.34) | 86.6 (5.04) | 90.3 (3.63) | 94.1 (4.20) |
| **Forehead** | | | | | |
| **Participant #** | **Day 0** | **Day 7** | **Day 14** | **Day 21** | **Day 28** |
| 1 | 66 | 69 | 77 | 82 | 86 |
| 2 | 76 | 82 | 86 | 89 | 90 |
| 3 | 69 | 73 | 82 | 86 | 88 |
| 4 | 63 | 68 | 73 | 76 | 84 |
| 5 | 52 | 58 | 67 | 71 | 73 |
| 6 | 52 | 57 | 64 | 73 | 76 |
| 7 | 42 | 44 | 53 | 58 | 60 |
| 8 | 69 | 82 | 83 | 87 | 89 |
| 9 | 53 | 61 | 68 | 74 | 74 |
| 10 | 67 | 76 | 80 | 86 | 89 |
| 11 | 74 | 79 | 84 | 87 | 89 |
| 12 | 70 | 78 | 81 | 85 | 89 |
| 13 | 61 | 64 | 69 | 73 | 76 |
| 14 | 72 | 76 | 81 | 84 | 87 |
| 15 | 56 | 59 | 65 | 71 | 76 |
| 16 | 57 | 62 | 68 | 84 | 86 |
| 17 | 51 | 55 | 66 | 72 | 75 |
| 18 | 54 | 58 | 67 | 71 | 73 |
| 19 | 75 | 78 | 81 | 86 | 88 |
| 20 | 60 | 66 | 74 | 79 | 83 |
| Mean (SD) | 62.0 (9.58) | 67.3 (10.53) | 73.5 (8.76) | 78.7 (8.13) | 81.6 (8.13) |

Table S5. Redness score (%, higher = less redness) during the study period.

| **Left cheek** | | | | | |
| --- | --- | --- | --- | --- | --- |
| **Participant #** | **Day 0** | **Day 7** | **Day 14** | **Day 21** | **Day 28** |
| 1 | 50 | 57 | 76 | 78 | 82 |
| 2 | 42 | 47 | 49 | 70 | 71 |
| 3 | 85 | 86 | 87 | 88 | 90 |
| 4 | 37 | 39 | 41 | 44 | 54 |
| 5 | 87 | 88 | 89 | 91 | 92 |
| 6 | 87 | 89 | 91 | 92 | 95 |
| 7 | 44 | 52 | 54 | 55 | 62 |
| 8 | 66 | 71 | 73 | 79 | 79 |
| 9 | 64 | 69 | 70 | 74 | 77 |
| 10 | 53 | 55 | 56 | 70 | 71 |
| 11 | 86 | 87 | 88 | 89 | 89 |
| 12 | 57 | 60 | 62 | 68 | 69 |
| 13 | 81 | 82 | 85 | 86 | 89 |
| 14 | 69 | 76 | 78 | 79 | 80 |
| 15 | 61 | 71 | 73 | 75 | 82 |
| 16 | 77 | 78 | 80 | 82 | 91 |
| 17 | 45 | 48 | 52 | 55 | 56 |
| 18 | 78 | 85 | 89 | 92 | 95 |
| 19 | 54 | 56 | 64 | 67 | 73 |
| 20 | 44 | 57 | 68 | 73 | 78 |
| Mean (SD) | 63.4 (17.02) | 67.7 (15.81) | 71.3 (15.13) | 75.4 (13.25) | 78.8 (12.30) |
| **Right cheek** | | | | | |
| **Participant #** | **Day 0** | **Day 7** | **Day 14** | **Day 21** | **Day 28** |
| 1 | 43 | 44 | 54 | 68 | 77 |
| 2 | 55 | 59 | 62 | 65 | 69 |
| 3 | 87 | 88 | 89 | 91 | 93 |
| 4 | 42 | 53 | 60 | 66 | 78 |
| 5 | 66 | 74 | 78 | 81 | 85 |
| 6 | 77 | 79 | 80 | 81 | 83 |
| 7 | 51 | 52 | 56 | 64 | 66 |
| 8 | 73 | 78 | 88 | 93 | 93 |
| 9 | 50 | 54 | 64 | 69 | 74 |
| 10 | 67 | 68 | 74 | 84 | 85 |
| 11 | 85 | 88 | 89 | 92 | 94 |
| 12 | 54 | 56 | 60 | 62 | 66 |
| 13 | 81 | 82 | 84 | 87 | 88 |
| 14 | 51 | 53 | 59 | 63 | 71 |
| 15 | 67 | 68 | 73 | 75 | 79 |
| 16 | 70 | 81 | 82 | 84 | 87 |
| 17 | 37 | 39 | 40 | 51 | 82 |
| 18 | 82 | 83 | 89 | 90 | 93 |
| 19 | 58 | 60 | 61 | 75 | 78 |
| 20 | 76 | 78 | 80 | 82 | 93 |
| Mean (SD) | 63.6 (15.28) | 66.9 (15.09) | 71.1 (14.26) | 76.2 (12.08) | 81.7 (9.29) |
| **Forehead** | | | | | |
| **Participant #** | **Day 0** | **Day 7** | **Day 14** | **Day 21** | **Day 28** |
| 1 | 62 | 65 | 68 | 75 | 91 |
| 2 | 72 | 74 | 77 | 78 | 84 |
| 3 | 88 | 89 | 90 | 91 | 94 |
| 4 | 43 | 47 | 60 | 62 | 65 |
| 5 | 82 | 83 | 86 | 87 | 89 |
| 6 | 84 | 88 | 90 | 92 | 94 |
| 7 | 41 | 55 | 56 | 61 | 67 |
| 8 | 77 | 79 | 85 | 85 | 87 |
| 9 | 82 | 85 | 86 | 87 | 91 |
| 10 | 53 | 66 | 67 | 75 | 78 |
| 11 | 88 | 89 | 90 | 91 | 93 |
| 12 | 74 | 75 | 76 | 79 | 81 |
| 13 | 73 | 77 | 80 | 90 | 92 |
| 14 | 60 | 67 | 78 | 81 | 85 |
| 15 | 70 | 80 | 84 | 88 | 90 |
| 16 | 81 | 89 | 90 | 92 | 93 |
| 17 | 58 | 59 | 62 | 63 | 76 |
| 18 | 72 | 80 | 85 | 92 | 92 |
| 19 | 68 | 70 | 77 | 81 | 84 |
| 20 | 65 | 69 | 77 | 88 | 91 |
| Mean (SD) | 69.7 (13.63) | 74.3 (11.98) | 78.2 (10.59) | 81.9 (10.22) | 85.9 (8.58) |

Table S6. Pores score (%, higher = less pores) during the study period.

| **Left cheek** | | | | | |
| --- | --- | --- | --- | --- | --- |
| **Participant #** | **Day 0** | **Day 7** | **Day 14** | **Day 21** | **Day 28** |
| 1 | 34 | 38 | 42 | 47 | 51 |
| 2 | 37 | 42 | 46 | 51 | 55 |
| 3 | 53 | 65 | 70 | 76 | 83 |
| 4 | 51 | 54 | 60 | 63 | 67 |
| 5 | 59 | 66 | 68 | 70 | 72 |
| 6 | 23 | 24 | 29 | 34 | 35 |
| 7 | 31 | 34 | 39 | 45 | 48 |
| 8 | 62 | 66 | 70 | 79 | 83 |
| 9 | 43 | 45 | 50 | 54 | 63 |
| 10 | 44 | 46 | 55 | 59 | 61 |
| 11 | 70 | 75 | 81 | 90 | 92 |
| 12 | 25 | 27 | 29 | 32 | 36 |
| 13 | 26 | 27 | 29 | 33 | 34 |
| 14 | 30 | 38 | 41 | 44 | 46 |
| 15 | 53 | 54 | 57 | 64 | 69 |
| 16 | 28 | 32 | 36 | 39 | 43 |
| 17 | 78 | 82 | 84 | 85 | 86 |
| 18 | 36 | 39 | 45 | 52 | 55 |
| 19 | 37 | 39 | 40 | 45 | 52 |
| 20 | 41 | 45 | 52 | 55 | 59 |
| Mean (SD) | 43.1 (15.55) | 46.9 (16.54) | 51.2 (16.73) | 55.9 (17.22) | 59.5 (17.4) |
| **Right cheek** | | | | | |
| **Participant #** | **Day 0** | **Day 7** | **Day 14** | **Day 21** | **Day 28** |
| 1 | 33 | 35 | 39 | 41 | 46 |
| 2 | 32 | 36 | 40 | 42 | 44 |
| 3 | 69 | 77 | 82 | 85 | 86 |
| 4 | 37 | 40 | 42 | 47 | 58 |
| 5 | 47 | 56 | 57 | 58 | 61 |
| 6 | 27 | 29 | 31 | 33 | 37 |
| 7 | 34 | 38 | 40 | 44 | 53 |
| 8 | 56 | 59 | 62 | 65 | 71 |
| 9 | 57 | 60 | 68 | 75 | 88 |
| 10 | 41 | 45 | 49 | 53 | 58 |
| 11 | 68 | 72 | 81 | 86 | 93 |
| 12 | 26 | 31 | 36 | 42 | 41 |
| 13 | 33 | 34 | 42 | 46 | 47 |
| 14 | 24 | 27 | 28 | 31 | 32 |
| 15 | 19 | 29 | 28 | 19 | 25 |
| 16 | 25 | 29 | 33 | 37 | 38 |
| 17 | 68 | 76 | 77 | 81 | 85 |
| 18 | 33 | 37 | 42 | 46 | 50 |
| 19 | 33 | 35 | 38 | 39 | 49 |
| 20 | 44 | 49 | 53 | 58 | 62 |
| Mean (SD) | 40.3 (15.58) | 44.7 (16.38) | 48.4 (17.26) | 51.4 (18.66) | 56.2 (19.59) |
| **Forehead** | | | | | |
| **Participant #** | **Day 0** | **Day 7** | **Day 14** | **Day 21** | **Day 28** |
| 1 | 32 | 35 | 41 | 43 | 46 |
| 2 | 30 | 33 | 37 | 41 | 40 |
| 3 | 46 | 50 | 62 | 66 | 72 |
| 4 | 33 | 35 | 36 | 37 | 46 |
| 5 | 32 | 36 | 40 | 42 | 45 |
| 6 | 22 | 25 | 28 | 32 | 33 |
| 7 | 31 | 35 | 37 | 41 | 45 |
| 8 | 51 | 54 | 55 | 59 | 62 |
| 9 | 53 | 58 | 62 | 75 | 83 |
| 10 | 43 | 47 | 48 | 52 | 57 |
| 11 | 57 | 65 | 67 | 72 | 74 |
| 12 | 23 | 26 | 28 | 31 | 35 |
| 13 | 31 | 32 | 38 | 39 | 44 |
| 14 | 29 | 31 | 35 | 37 | 43 |
| 15 | 45 | 51 | 57 | 61 | 63 |
| 16 | 23 | 26 | 30 | 32 | 33 |
| 17 | 66 | 74 | 77 | 83 | 87 |
| 18 | 31 | 34 | 38 | 42 | 45 |
| 19 | 59 | 60 | 61 | 68 | 75 |
| 20 | 45 | 48 | 50 | 52 | 60 |
| Mean (SD) | 39.1 (13.05) | 42.8 (14.18) | 46.4 (14.21) | 50.3 (15.88) | 54.4 (16.72) |
