## Supplementary Figures for "Evaluation of Effects on Skin Quality of a *Centella asiatica* Extracellular Vesicle-based Skin Care Formulation: A 28-Day Facial Skin Quality Study"

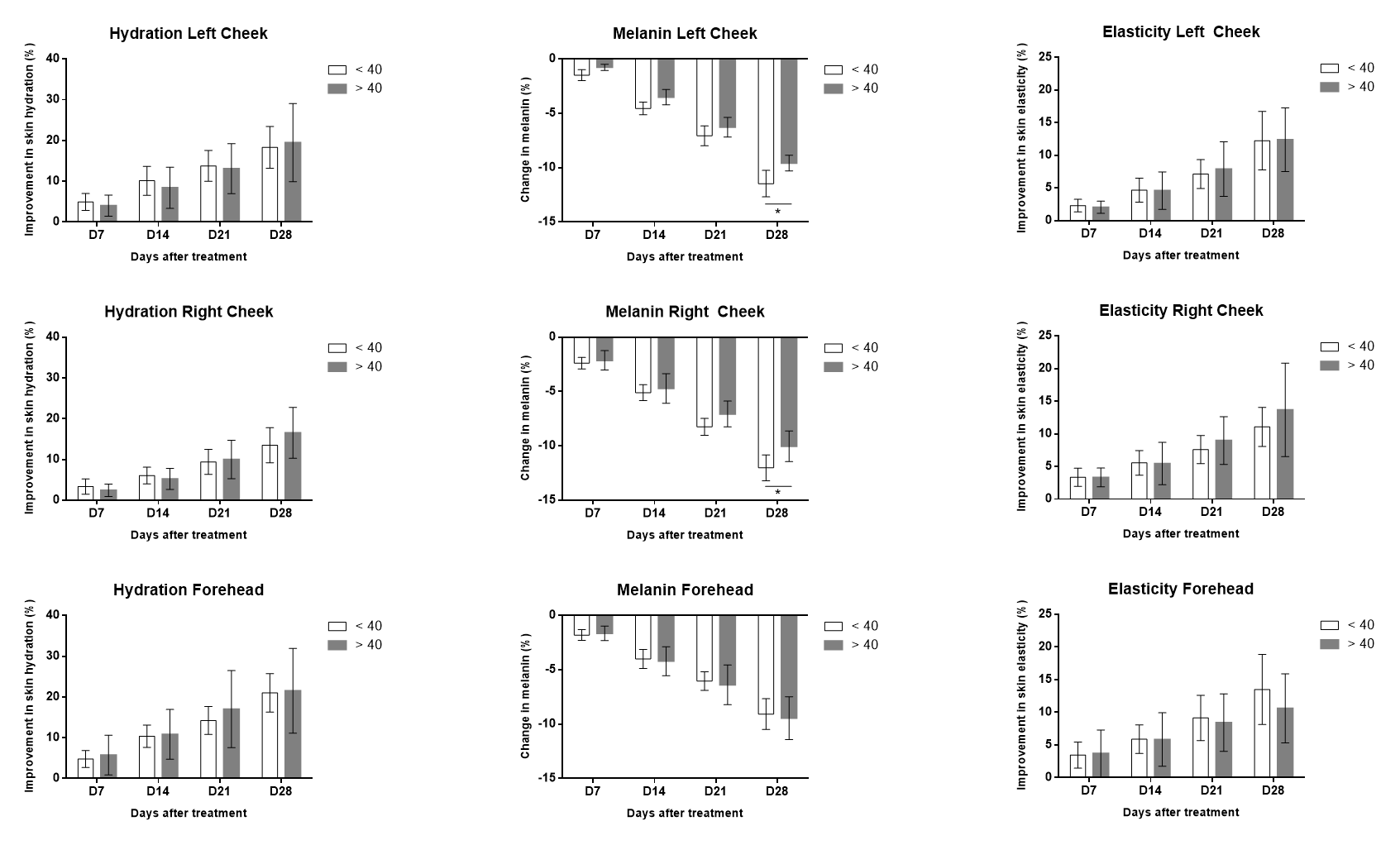

Figure S1. Change in skin hydration (left column), melanin (middle column), and elasticity (right column) at 7, 14, 21, and 28 days after treatment of test product in left cheek (top row), right cheek (middle row), and forehead (bottom row) facial regions. Results are shown as bars representing means grouped by participants less than age of 40 years (white) and greater than age of 40 years (gray) with error bars representing 95% confidence interval of the mean. Statistical significances between means of participants in the two age groups at a single time point. * P<0.05

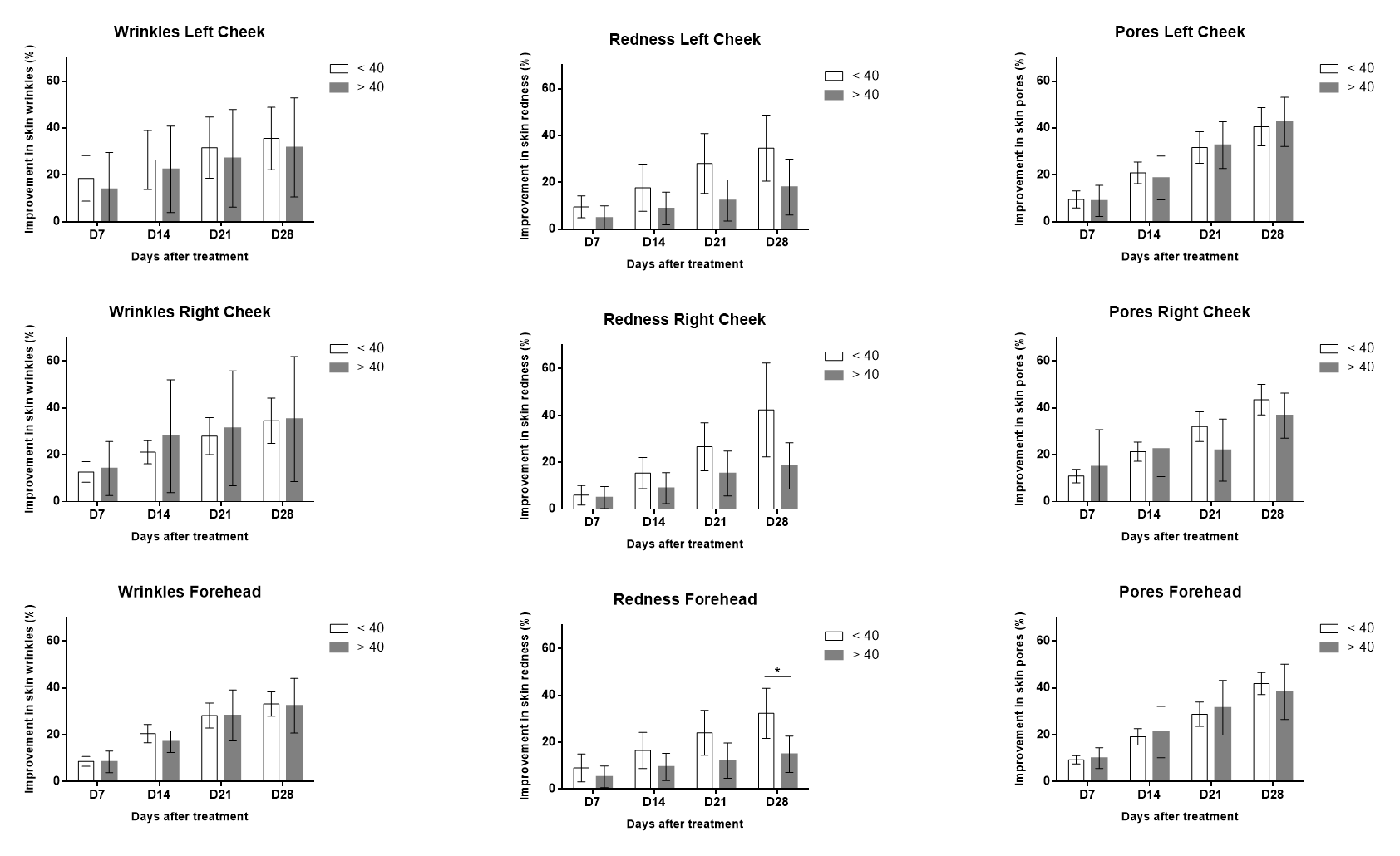

Figure S2. Change in skin wrinkles (left column), redness (middle column), and pores (right column) at 7, 14, 21, and 28 days after treatment of test product in left cheek (top row), right cheek (middle row), and forehead (bottom row) facial regions. Results are shown as bars representing means grouped by participants less than age of 40 years (white) and greater than age of 40 years (gray) with error bars representing 95% confidence interval of the mean. Statistical significances between means of participants in the two age groups at a single time point. * P<0.05
